# Survival and Predictor of Thrombocytopenic Neonatal Death in Public Hospitals of Addis Ababa, Ethiopia, 2025: Multicenter Prospective Follow Up Study

**DOI:** 10.1101/2025.08.01.25332567

**Authors:** Yohannes Godie Ashebir, Fikirte Kassaye, Teklu Assefa, Taddle Abate, Tiruye Menshaw, Misrak Tafese, Misgana Hirpha, Asalef Endazanaw

## Abstract

**Background:** Globally, thrombocytopenia is one of the most common hematologic conditions seen in ill neonates. In countries with limited resources, like Ethiopia, it is a serious concern. Because the burden of thrombocytopenia is so great, generating updates evidence on predictors of mortality and survival status is vital to fight it. However, the problem is not well investigated in Addis Ababa. Therefore, this study aimed to assess survival and predictor of thrombocytopenic neonatal death in Public Hospitals, Addis Ababa, Ethiopia, 2024/2025.

**Methods and Materials:** A prospective follow-up study was done among a total of 350 neonates from March 20, 2025, to April 30, 2025, in Addis Ababa public hospitals. All thrombocytopenic neonates that meet the inclusion criteria were chosen as study participants. Data were collected using the Kobo Tool through direct observation and review of maternal and neonatal charts. After export to an Excel spreadsheet, data cleaning and recoding were performed using SPSS version 26, followed by statistical analysis using STATA version 17. The Kaplan-Meier failure curve was used to demonstrate the pattern of death, estimate the chance of death, and compare failure curves. Collinearity, Schoenfeld residual, and log-rank tests were performed. The Cox proportional hazards model was fitted with global test result of 0.7882. Finally, the findings were presented both descriptively and analytically.

**Results:** In this study, the overall magnitude of thrombocytopenic neonatal death was 14.1% (95% CI: 10.4–18.1), with an incidence rate of 13.04/1000 (95% CI: 0.009–0.017) neonate-days.

The restricted mean time to death in this study was 23.36 days (95% CI: 22.23–24.50). Being born to a mother with severe preeclampsia (AHR = 3.84; 95% CI: 1.78–8.26), very low birth weight (<1499g) (AHR = 3.67; 95% CI: 1.14–11.80), perinatal asphyxia (AHR = 2.76; 95% CI: 1.32–5.79), necrotizing enterocolitis (AHR = 2.45; 95% CI: 1.14–5.31), and delayed initiation of feeding (AHR = 3.37; 95% CI: 1.10–10.29) were the identified predictors of mortality.

**Conclusion and recommendation:** In this study, a high burden of thrombocytopenic neonatal death. Early detection and treatment of high-risk conditions like severe preeclampsia, very low birth weight, perinatal asphyxia, and necrotizing enterocolitis should be the main goal of efforts to lower thrombocytopenic neonatal mortality. Furthermore, prompt neonatal feeding initiation ought to be given top priority.

## INTRODUCTION

A neonate refers to a newborn infant in the first 28 days of life (1,2). Addressing neonatal mortality is an important priority for improving the health and well-being of newborns (3). Neonates are particularly vulnerable to sepsis, temperature instability, respiratory distress, hematologic problems, and feeding difficulties(4). Among those hematological problems thrombocytopenia is the frequent disorder commonly seen in neonates (5). Though, the impact of thrombocytopenia is high on neonates with intrauterine growth restriction, polycythemia, perinatal asphyxia, congenital infections,disseminated intravascular coagulation and associated with systemic diseases sepsis, hypoxia, and necrotizing enterocolitis (6), prematurity is an important risk factor for thrombocytopenia (7).

Neonatal thrombocytopenia is one of the problems of neonates that contributes high mortality of neonates with a high risk of bleeding (6). Several studies have found that incidences of thrombocytopenia among neonates are related with in Austria 56% (8), Pakistan,56.2% (9), Tunisia 63% (10), Sri Lanka, 58.2% (11), and India, 63.33% (12) according to several research.. Thrombocytopenia occurs in 220–350/1000 newborns admitted to a neonatal intensive care unit and up to 700–800/1000 in very low-birth-weight and preterm neonates admitted to the neonatal intensive care unit who require intensive care (13). It is one of the most common hematologic disorders in neonatal intensive care units, occurring in up to a quarter of admissions, and leads to a high risk of bleeding and mortality (14).

Thrombocytopenia is the most common bleeding disorder in the newborn, with 20% to 35% of all NICU admissions and 70% of infants born at less than 1,000g having the diagnosis during their NICU (15). Evidences show that thrombocytopenia 282 (76%) were preterm born(16) and at least 30 (75%) neonates were still thrombocytopenic at death (16). Thrombocytopenia in preterm neonates with birth weight <1000 g and <750g is reported to occur in up to 75 and 90% of neonates, respectively (17). Thrombocytopenia has been independently related to mortality and major morbidities (18) by contributing to high incidence of intracranial hemorrhage and bleeding (19).

The incidence of thrombocytopenia is inversely proportional to the gestational age, and it represents a risk factor for poor neonatal outcomes and bleeding from thrombocytopenia is a potentially life-threatening (5). It is a serious disorder affecting 15-40% of critically ill neonates (20). Thrombocytopenia has overwhelmed effect on neonates having sepsis and mortality is significantly high in neonatal sepsis along with thrombocytopenia (21). Updated evidence indicates that thrombocytopenia and platelet dysfunction contribute to failure of spontaneous and pharmacological patent ductus arteries closure in preterm infants (22). Thrombocytopenia in preterm neonates with necrotizing enterocolitis increase the risk of complications and mortality 50 to 95% (23,24). In sub-Sahara African countries, about 90% of cases of severe thrombocytopenia presenting after the first few days of life are due to late-onset bacterial sepsis, necrotizing enterocolitis, or both (25).

The symptoms of thrombocytopenia in neonate include bruising of skin (petechial), bleeding in other body systems, intravenous site bleeding and yellow skin and eye color (jaundice) because of bruising (26). Platelet transfusion is a specific treatment for thrombocytopenia (6). Platelet transfused neonates with gestational age <28 weeks had lower birth weights, were more often severely thrombocytopenic, and died more frequently than infants of a similar gestational age who were not transfused (27). The early diagnosis of neonatal thrombocytopenia and assessment of the underlying primary pathologic process play an important role in reducing the risk of life-threatening complications of neonatal thrombocytopenia (7).

In Ethiopia, neonatal thrombocytopenia is found to be prevalent which ranges from 49.3% to 66% and its severity is high on preterm neonates with respiratory distress, early onset neonatal sepsis and necrotizing enterocolitis (28–30). Though thrombocytopenia is prevalent it is often overlooked assuming it will resolve spontaneously (31). Despite efforts by the federal ministry of health and other partners to reduce newborn death, the rate fell from 39 to 29 between 2005 and 2016, but has since increased to 33, according to the 2019 Ethiopian demographic health survey (EDHS) report (32,33). This could be related to preventable or treatable causes of neonatal hematologic abnormalities, such as thrombocytopenia. As a result, identifying predictors of thrombocytopenic neonatal death and preventing, identifying, and monitoring this condition early on may improve neonatal outcomes. The purpose of this study is to assess survival and predictors of thrombocytopenic neonatal deaths in public hospitals of Addis Ababa, Ethiopia, 2024/2025.

## MATERIALS AND METHODS

### Study area, Study Design and Period

This study was conducted in Addis Ababa public hospitals. From the total of 12 hospitals in the city, six hospitals were selected by lottery methods. These selected hospitals are Alert Comprehensive Specialized Hospital, Gandhi Memorial Hospital, Menelik II Comprehensive Specialized Hospital, Yekatit 12 Hospital Medical College, St. Peter Specialized Hospital, and Zewditu Memorial Hospital. A prospective cohort follow-up study was conducted from March 20, 2025, to May 30, 2025.

Institutional based Cross-sectional study design was employed. The study was conducted in East Gojjam Zone Public Hospitals. In Addis Ababa Zone there were 4 Hospitals (Debre Markos Referral Hospital, Lumamie Hospital, Bichena Hospiatl and Shegaw Motta Hospital) with a total of 284 nurses during this study. By simple random sampling 181 nurses were included in the study as the study participant.

### Source and study Populations

Source of population, all thrombocytopenic neonates were found in neonatal intensive care units of Addis Ababa public hospitals. In a study population, all thrombocytopenic neonates were found in selected Addis Ababa public hospitals’ NICUs from March 20, 2025, to April 30, 2025.

### Inclusion and Exclusion Criteria

Inclusion criteria, neonates with thrombocytopenia in selected NICUs of Addis Ababa public hospitals during the study period were included. Thrombocytopenic neonates diagnosed with neural tube defect, abdominal wall defect, or congenital cardiac disease and whose mothers were critically ill or unable to give informed consent were excluded. Thrombocytopenic neonates who have no indexed attendant or those who are abandoned neonates were excluded.

### Sample Size Determination

The sample size is calculated using a formula designed for survival analysis in STATA Version 17 statistical software, taking into account significant determinants of time to death in neonates with thrombocytopenia (34). Based on this literature, the following are calculated.

The survival sample size was determined using STATA version 17 with the formula N = E/P (E) by the proportion from prior studies (Table 1).

**Table 1:** sample size calculation for neonates with thrombocytopenia.

| Proportion Death (P) | Estimated Hazard ratio | Error probability | Power | Proportion of withdrawals | N |
| --- | --- | --- | --- | --- | --- |
| 43.2%(0.432) (Farag *et al.*, 2024) | 0.5 | 0.05 | 80% | 0.1 | 169 |
| **20.8%(0.208)**<br>(Thrombocytopenia *et al.*, ) | **0.5** | **0.05** | **80%** | **0.1** | **350** |
| 25.3%(0.253)(Peng *et al.*, 2022) | 0.5 | 0.05 | 80% | 0.1 | 287 |

For this, the largest sample size (350) is selected to be the final sample size of the study. As far as the searching trial to bring hazard ratio for the predictors, there was no study with a predictor reported by hazard ratio. Therefore, the estimated hazard ratio (0.5) was used to calculate sample size.

### Sampling Procedure

Samples were recruited from each selected hospital prospectively starting from the time of diagnosis of neonatal thrombocytopenia. Three-month average baseline neonates diagnosed with thrombocytopenia data were taken from HMIS from each selected hospital. Based on the data, Alert Comprehensive Specialized Hospital (72), Gandhi Memorial Hospital (74), Menelik II Comprehensive Specialized Hospital (56), Yekatit 12 Hospital Medical College (71), St. Peter Specialized Hospital (67), and Zewditu Memorial Hospital have an average monthly admission of 62. From this, the value of k was 402/350=1.2. Therefore, by using the consecutive sampling technique, from each neonate diagnosed with thrombocytopenia that fully fulfills the inclusion criteria, study participants were selected until the required sample size is achieved in each hospital. If the recruited sample index mother refuses to participate, the next admission was employed.

### Study Variables

#### Dependent Variable

Time to thrombocytopenic neonatal death

### Independent Variables

#### Neonatal and maternal-related predictors

sex, maternal age, educational status, weight of neonate, age of neonate, and date of admission and discharge.

#### Obstetric-related predictors

multiple pregnancies, PROM, mode of delivery, preeclampsia, abruptio placentae, ANC follow-up, steroid administration, hypertension, DM, and HIV/AIDS.

#### Clinical variables

PNA, RD, sepsis, jaundice, hypothermia, hypoglycemia, NEC

#### Treatment and health service-related variables

platelet transfusion, antibiotics, CPAP, KMC, and nurse-to-patient ratio.

#### Operational Definitions/ Definition of Terms

Death: Neonate diagnosed with thrombocytopenic who were die during the follow-up within 28 days after birth Censored: thrombocytopenic neonates who left the follow up without event (transferred to another institution, left against medical advice, recovered from thrombocytopenia discharged and neonates stays more than 28 days)

Event: thrombocytopenic neonate who were die during the follow-up Time to death: time at which the neonates died during the follow up Follow up time period: Time from recruiting up to either the study subjects died or censored

### Data Collection Tools and Procedures

To collect the data for this study, the tool was adapted from different studies (14,21,28,35) and adapted by expert neonatal nurses and neonatologists. The tool is prepared in English and translated to Amharic and then re-translated to English. Maternal-related data were collected by using the mother’s medical charts. Neonatal data were collected from medical records and observations. The data collection tool includes maternal, neonatal, clinical, and treatment-related predictors by considering time to thrombocytopenic neonatal death as an outcome variable. The data were collected by twelve neonatal nurses. The completeness of each record was checked daily soon after collecting the data. The follow-up was at least two times per 24 hours. If the study subject is referred to another institution that is in the study area, the follow-up continues. Otherwise, the study subject was considered censored.

### Data Quality Control and Management

The tools were externally validated by senior researchers, and a pretest was conducted on 35 thrombocytopenic neonates at Ras Desta Damtew Hospital. Data collectors and supervisors received ten days of training on data collection tools and procedures. Supervisors closely monitored the data collection process. The research team and supervisors were checking the completeness of the data for each study subject outcome.

## Data Analysis

Data were collected using KoboToolbox, and finally they were exported to Excel. After cleaning and organizing, it was exported to SPSS version 26 for recoding and further cleaning. For the sake of analysis, it was transported to STATA version 17. Before regression, the outcomes were dichotomized as death or event (“1”) and censored or non-event (“0”). The time to death was calculated in days using the time interval between the time of diagnosis and the time of outcome. Multi-collinearity for variable similarities using VIF (1.18), a log-rank test to see the survival difference between or among the categories. Bi-variable Cox regression was first fitted, and those independent variables having the determined p-value were included in the multi-variable Cox regression analysis, which was fitted at a 5% level of significance to determine the net effect of each explanatory predictor on the outcome variable (HR with its 95% confidence interval and p-values was used to measure strength of association and identify statistical significance). P-values < 0.05 in multivariable Cox proportional hazard regression were considered statistically significant predictors. Model fitness was tested by using Schoenfeld residuals test for proportionality assumption of each covariate and overall model of the stratified cox proportional hazard. Based on this test, the model was fitted with p-value of > 0.05 and global test result of 0.7882. Finally, the results of the study were presented with text, tables, and figures.

## RESULTS

### Socio-demographic predictors of the study participants

A total of 350 neonates and their index mothers were included with the response rate of 347(99.1%). The mean age of the mother was 28.7 ± 4.5 SD years. Indexed mothers’ marital status profile indicates that, majority of mothers were married 326(93.9%). Regarding educational status, most mothers had completed secondary education 108(31.1%). In this study most of the mother’s occupation were housewives 115(33.1%). (Table 2)

**Table 2:**
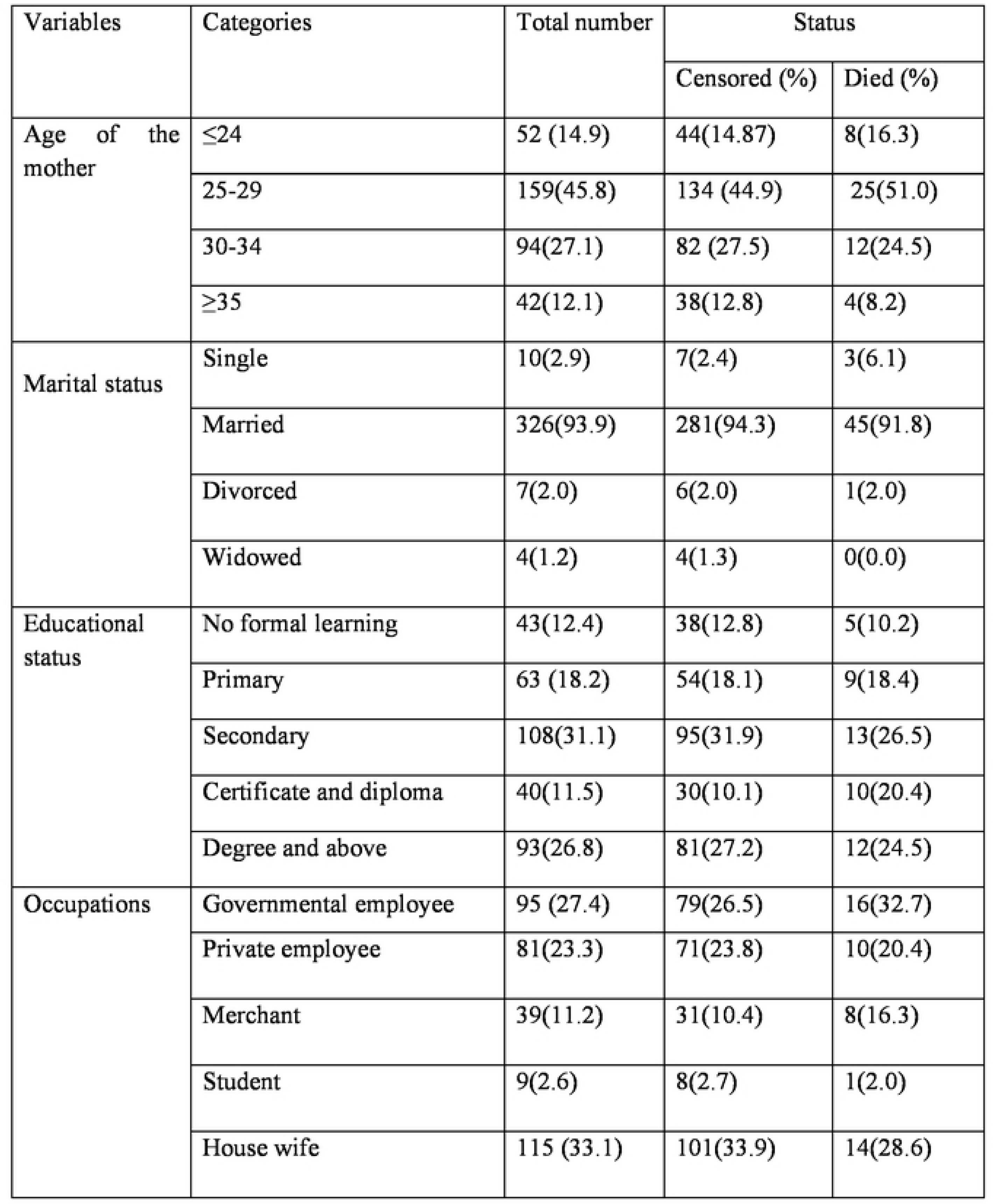
Socio-demographic characteristics of neonates and their index mother for the study of survival and predictors of thrombocytopenic neonatal death in public hospitals Addis Ababa, Ethiopia, 2025 (n=347)

### Maternal obstetric and medical related predictors

Out of the total study participants, 339(97.7%) had ANC follow-up, of whom most 316(86.5%) of them had four or more antenatal visit. Majority of the pregnancies were singleton 320(92.2%). Mode of delivery, half 176(50.7%) of mothers delivered through spontaneous vaginal delivery (SVD), 161(40.4%) by cesarean section (CS), and 10(2.9%) through instrumental delivery. Nearly all deliveries 342(98.6%) occurred in health facilities, while only 5(1.4%) took place at home. Among the facility deliveries, most 279(81.6%) were in hospitals and 63(18.4%) in health centers. About 67(19.3%) of neonates were referred from other institutions. More than half of the mothers 183(52.7%) experienced at least one obstetric complication during pregnancy or labor. Among these complications, severe preeclampsia was stated 49(14.1%), PROM 51(14.7%), APH 23(6.9%), and GDM 25(7.2%). (Table 3)

**Table 3:**
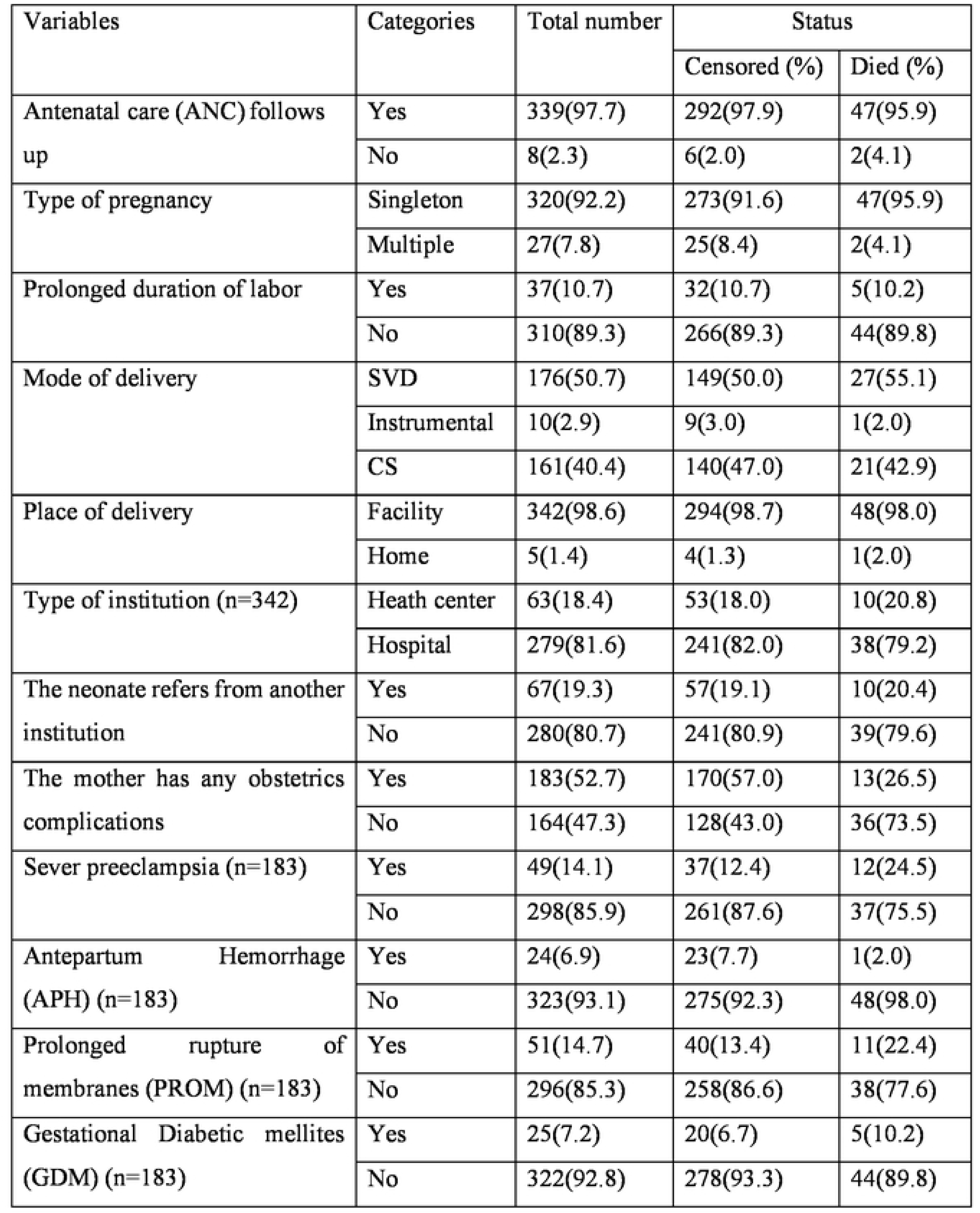
Maternal obstetrics, and medical related characteristics for the study of survival and predictors of thrombocytopenic neonatal death in public hospitals Addis Ababa, Ethiopia, 2025 (n=347)

### Neonatal demographical related predictors

Out of the total neonates included in the study, more than half were females 180 (51.9%). The age at admission showed that more than half 187 (53.9%) of neonates were admitted after one hour of birth. Regarding gestational age, preterm neonates (<36+6 weeks) represented 148 (42.7%). In terms of birth weight, nearly one-fourth 85(24.5%) of the neonates were very low birth weight neonates (<1499gram). Regarding weight-for-gestational-age, most neonates were appropriate for gestational age (AGA) 282 (81.3%). Regarding APGAR scores, at the 1st minute, nearly three-fourths, 241 (70.5%), of neonates scored between 7 and 8, and at the 5^th^ minute, most 301 (88.3%) of neonates scored 7 and 8. (Table 4)

**Table 4:**
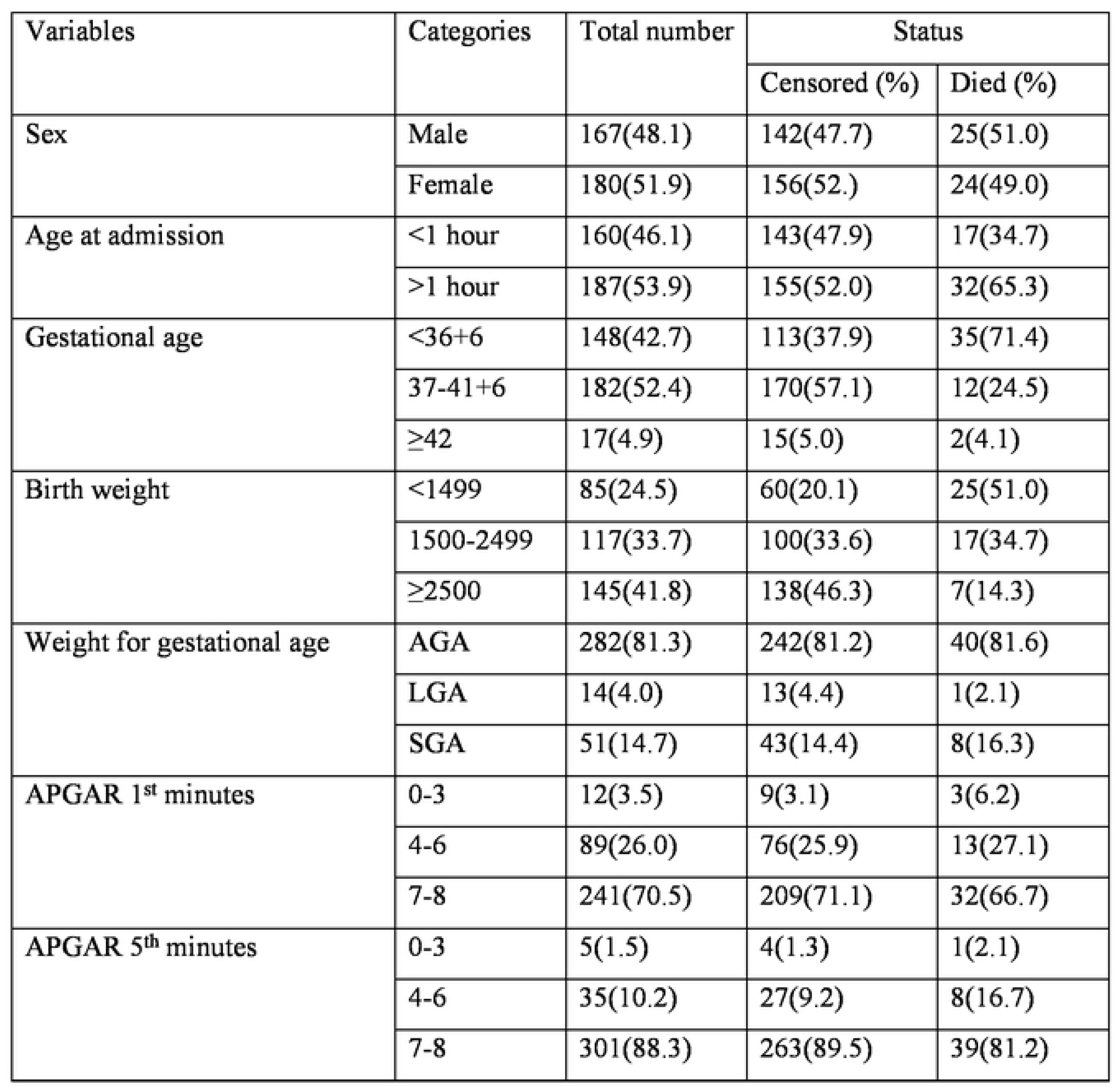
Neonatal demographical related characteristics for the study of survival and predictors of thrombocytopenic neonatal death in public hospitals Addis Ababa, Ethiopia, 2025 (n=347)

### Health service-related predictors

In this study, during the daytime, more than half, 183 (52.7%), of neonates were in rooms where the nurse-to-patient ratio was greater than 1:2. Similarly, during the nighttime, nearly three-fourths (249, or 71.7%) of neonates were cared for under a nurse-to-patient ratio of greater than 1:2. About physician follow-up, most 279 (80.4%) of the neonates were visited by a pediatrician or neonatologist twice per day. Most 304 (87.6%) of neonates had their vital signs checked four times daily. More than two-thirds, 248 (71.5%) of the neonates, CBC machine was always functional. Most 294 (84.7%) of neonates received regular nursing rounds. (Table 5)

**Table 5:**
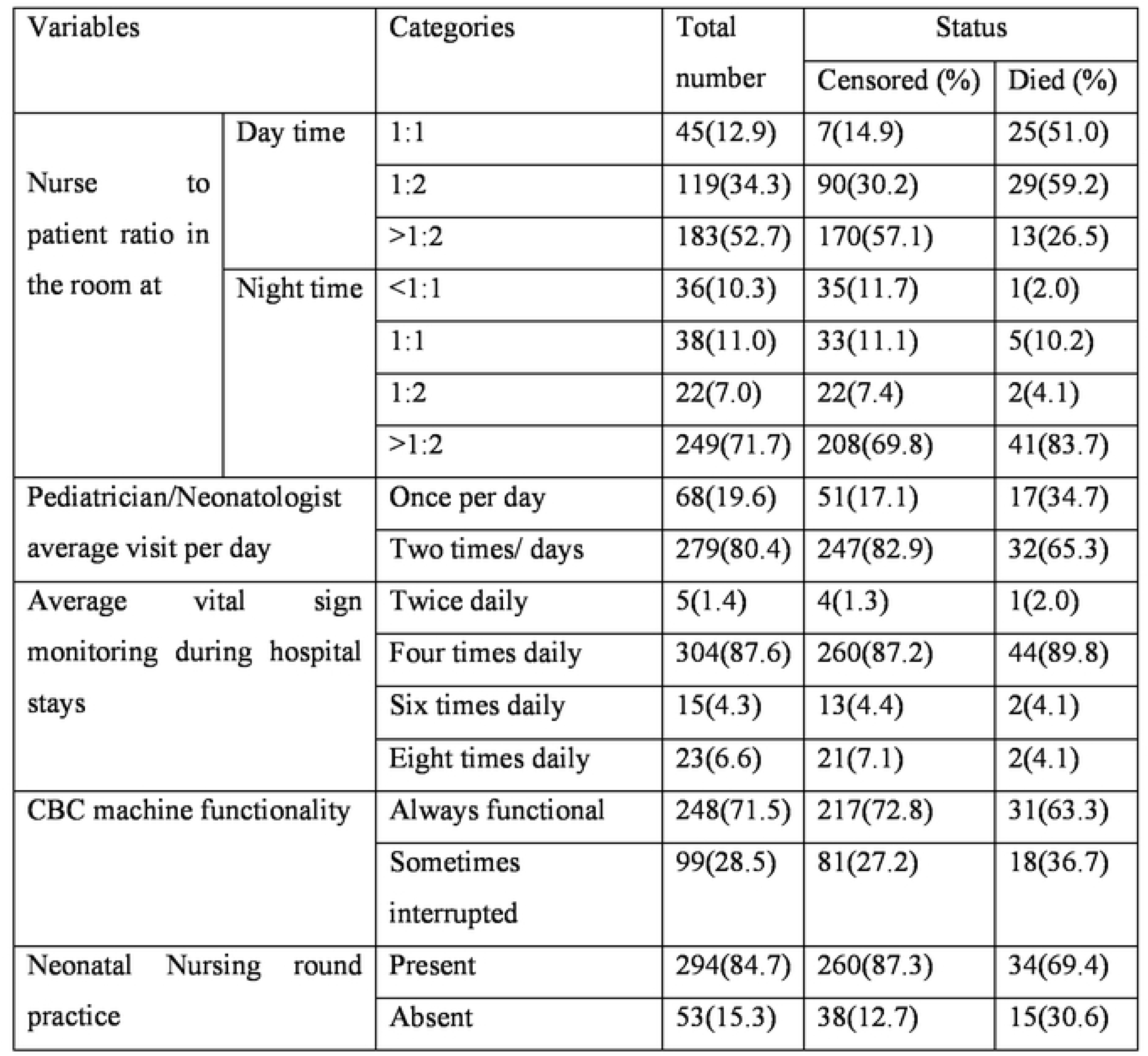
Health service-related characteristics for the study of survival and predictors of thrombocytopenic neonatal death in public hospitals Addis Ababa, Ethiopia, 2025 (n=347)

### Neonatal admission problem

Among the thrombocytopenic neonates admitted to the NICU, hypothermia was the most common admission problem, accounting for 281(31.2%), Respiratory distress (RD) 232(25.7%), Early-onset neonatal sepsis (EONS) 218(24.2%), Perinatal asphyxia (PNA) 76(8.4%), and others admission problem, including meningitis, jaundice, meconium aspiration syndrome (MAS), and hyperthermia 94(10.4%). (Figure 1)

**Figure 1:**
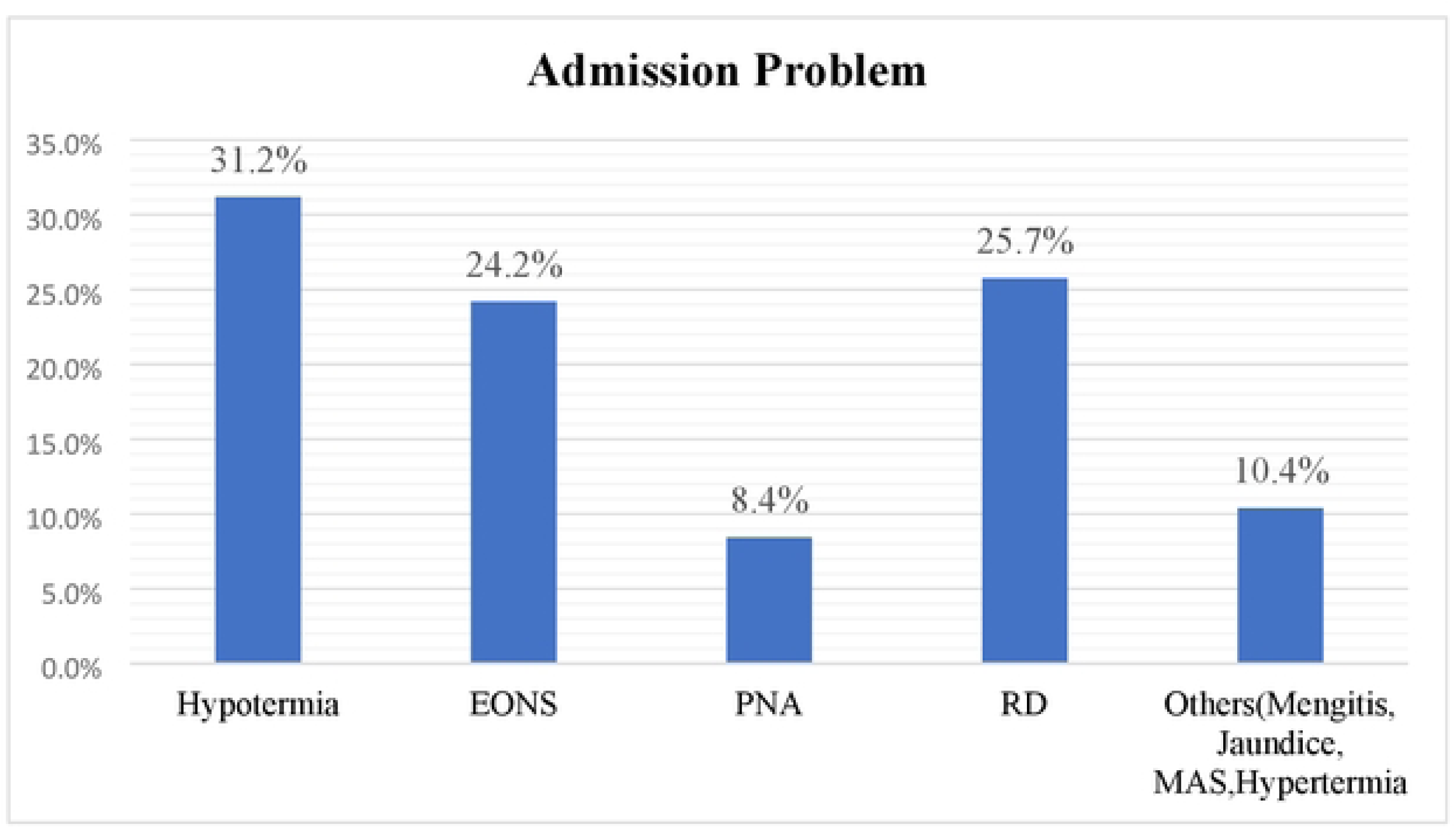
Neonatal admission problems for the study of survival and predictors of thrombocytopenic neonatal death in public hospitals Addis Ababa, Ethiopia, 2025 (n=347)

### Neonatal medical problem and treatment related predictors

Among the neonates admitted to the NICUs, nearly half, 164 (47.2%), had mild thrombocytopenia, while 148(42.7%) had moderate and 35(10.1%) had severe thrombocytopenia. Hospital-acquired infections (HAI) were diagnosed 151(43.5%), neonatal hyperbilirubinemia (NHB) 167(48.1%), apnea 27(7.8%), and necrotizing enterocolitis (NEC) 66 (19.0%). In terms of feeding initiation, only 126(36.8%) of neonates began feeding within the first 24 hours of life. Half of 167(50.2%) of neonate’s transfusion platelets. Of the transfused neonates, 109(65.3%) received transfusion within 4 hours of diagnosis. Regarding the age of platelet transfusion, most 147(88.0%) of the received platelets were less than 7 days old. The time between receiving lab results and transfusion, nearly half, 80(48%), of the study participants were transfused within one hour, and nearly one-third, 51(30.5%), of the neonates received other blood components. (Table 6)

**Table 6:**
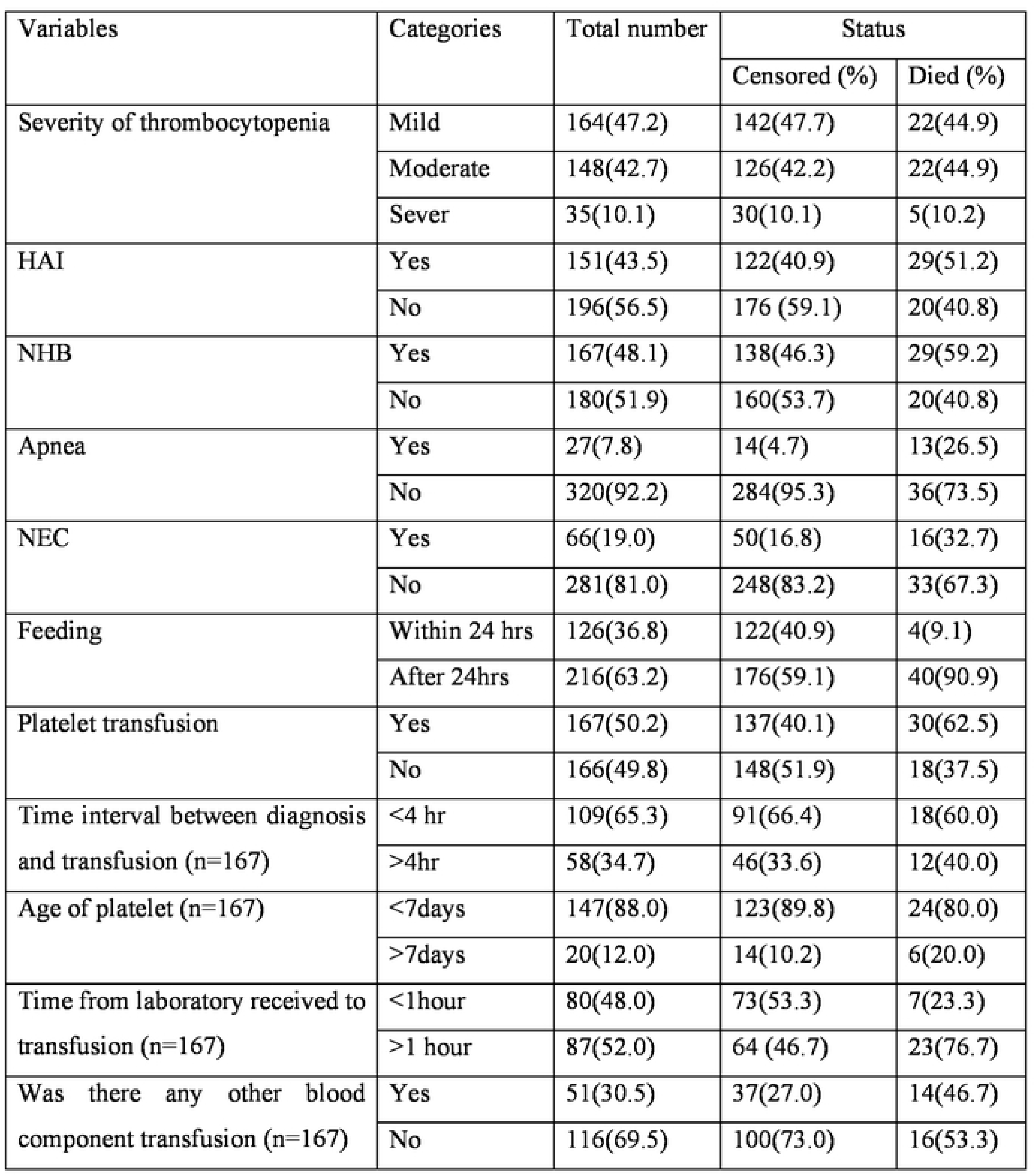
Neonatal medical problem and treatment related predictors for the study of survival and predictors of thrombocytopenic neonatal death in public hospitals Addis Ababa, Ethiopia, 2025.

In this study, among neonates diagnosed with thrombocytopenia, the most frequently administered treatments were antibiotics 323 (31.6%), oxygen therapy 248 (24.2%), intravenous (IV) fluid administration 219 (21.4%), phototherapy 152 (14.9%), whole blood transfusions 36 (3.5%), and other neonatal treatments such as Kangaroo Mother Care (KMC) and wound care were administered to 45 (4.4%) of the neonates. (Figure 2)

**Figure 2:**
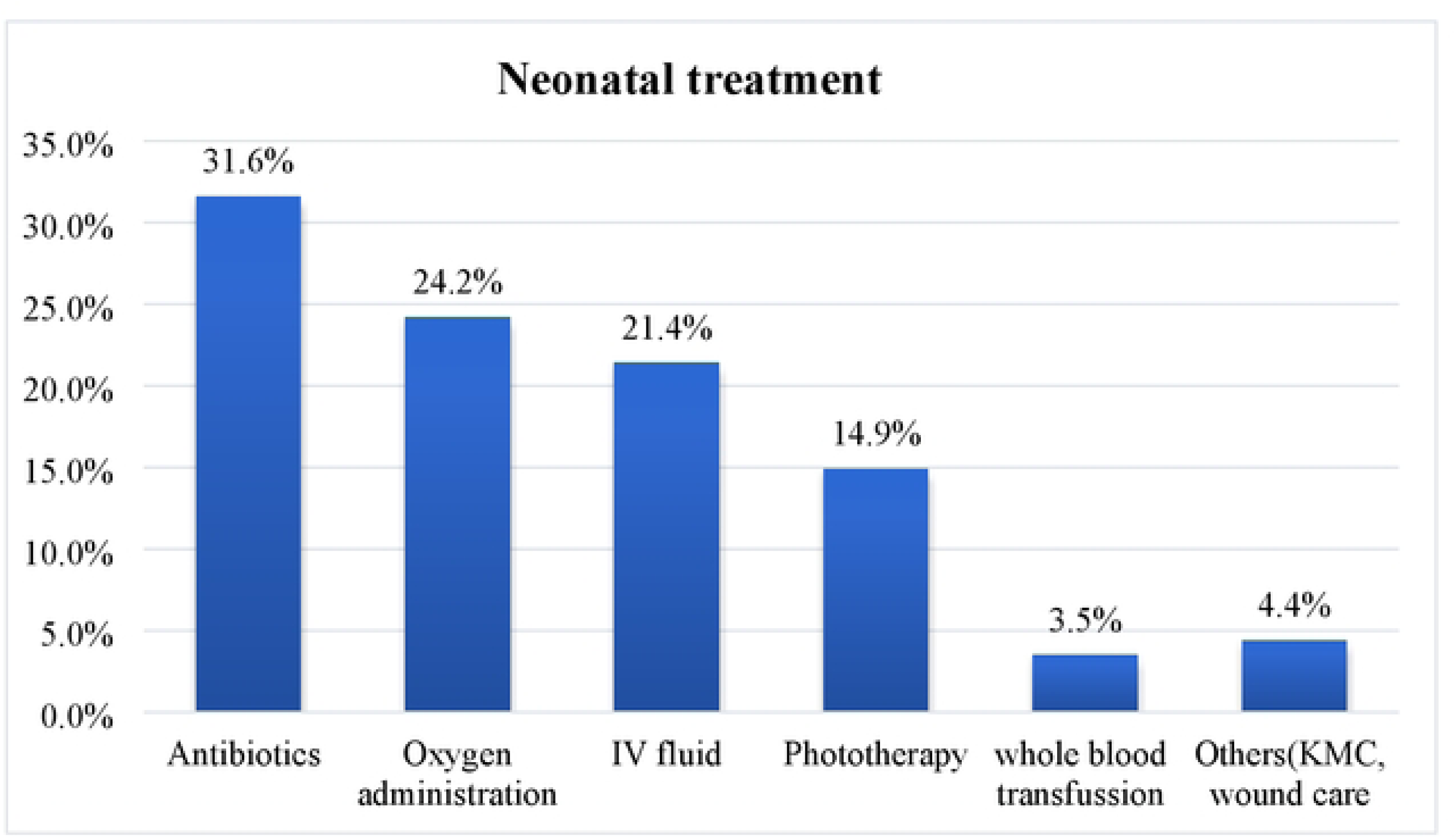
Neonatal treat1nent related factors for the study of survival and predictors of thro1nbocytopenic neonatal death in public hospitals Addis Ababa, Ethiopia, 2025 (n=347)

### Overall outcome of thrombocytopenic neonatal death

In this study a total of 347 neonates were followed for up to 28 days of age starting from admission up to the occurrence of outcome. Among those thrombocytopenic neonates 49(14.1%) of died. The most common immediate cause of neonatal death was cardiorespiratory failure 19 (38.8%), which was followed by multi-organ failure 15 (30.6%) and overwhelming sepsis 7 (14.3%). (Figure 3).

**Figure 3:**
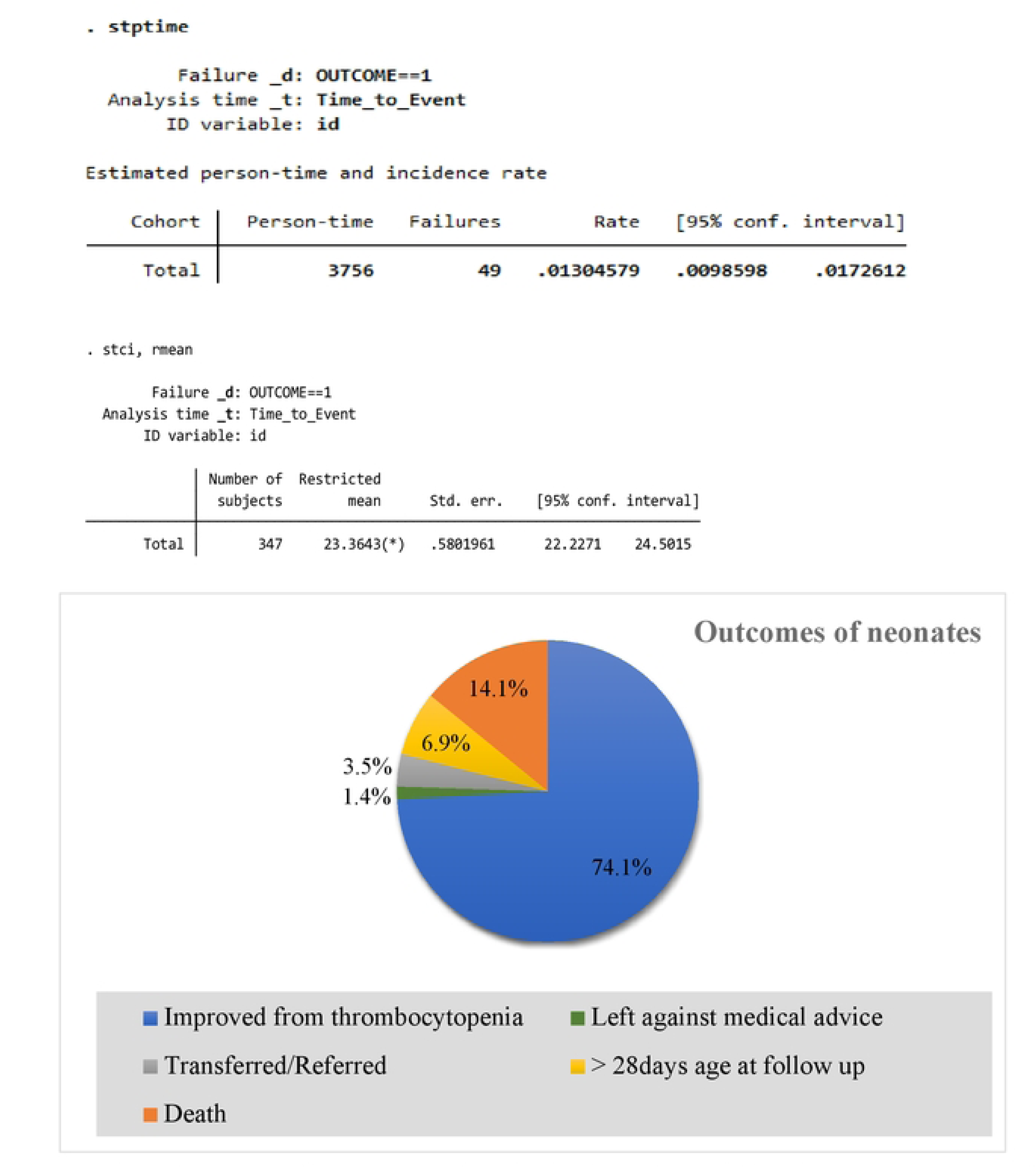
Over all outcomes for the study of survival and predictors ofthron1bocytopenic neonatal death in public hospitals Addis Ababa, Ethiopia, 2025 (n=347)

The overall proportion of death was 14.1% (95% CI: 10.4–18.1) with a person-day incidence rate of 1.30% (95% CI: 0.009–0.017), and the total analysis time was 3,756 person-days. The restricted mean time to death was 23.36 days (95% CI: 22.23–24.50), with a standard error of 0.58019.

### Overall Distribution of thrombocytopenic neonatal death

#### The Kaplan-Meier failure estimate curve provides an overall estimate of the time to death among thrombocytopenic neonates. The curve indicates a gradual increase in the failure probability (death) over the length of hospital stay, with significant steps occurring up to 28 days, after which the curve plateaus before reaching 50%, suggesting most deaths have occurred by this point. The number at risk decreases over time, reflecting neonates who either yielded to the condition or were censored, providing a clear visualization of survival probability declining as hospital stay extends. (Figure 4)

The log rank test results showed that there is no survival difference among thrombocytopenic neonates’ with and those without NEC with p-value of 0.0001. (Figure **5**). The log rank test result indicates that there is no a significant survival difference among thrombocytopenic neonates each category of gestational age with p-value of 0.0560 (Figure 6).

**Figure 4:**
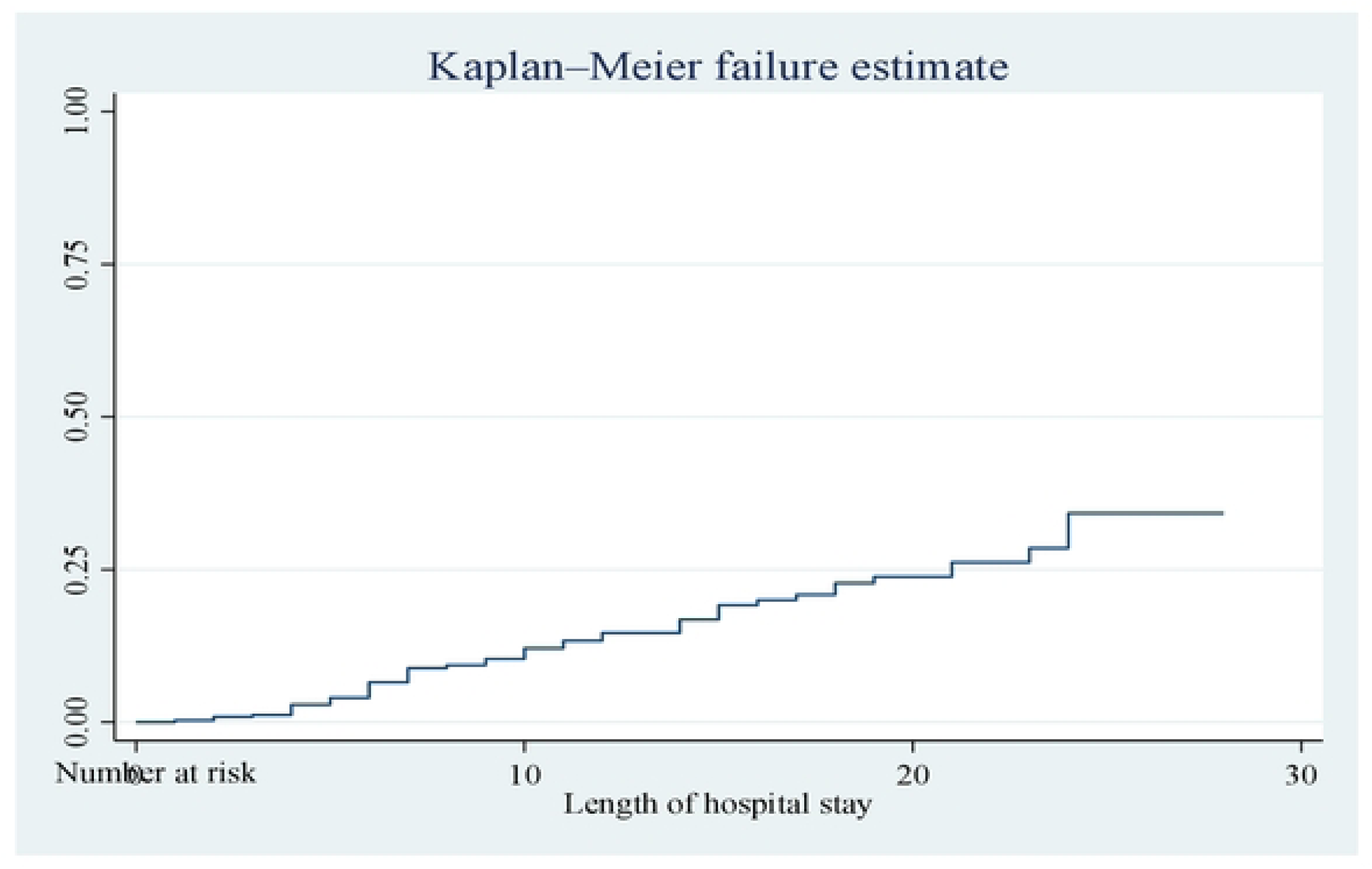
Over all Kaplan-Meier failure estimate of time to death among thrombocytopenic neonates admitted to NICU of Addis Ababa public hospitals, Ethiopia, 2025

**Figure 5:**
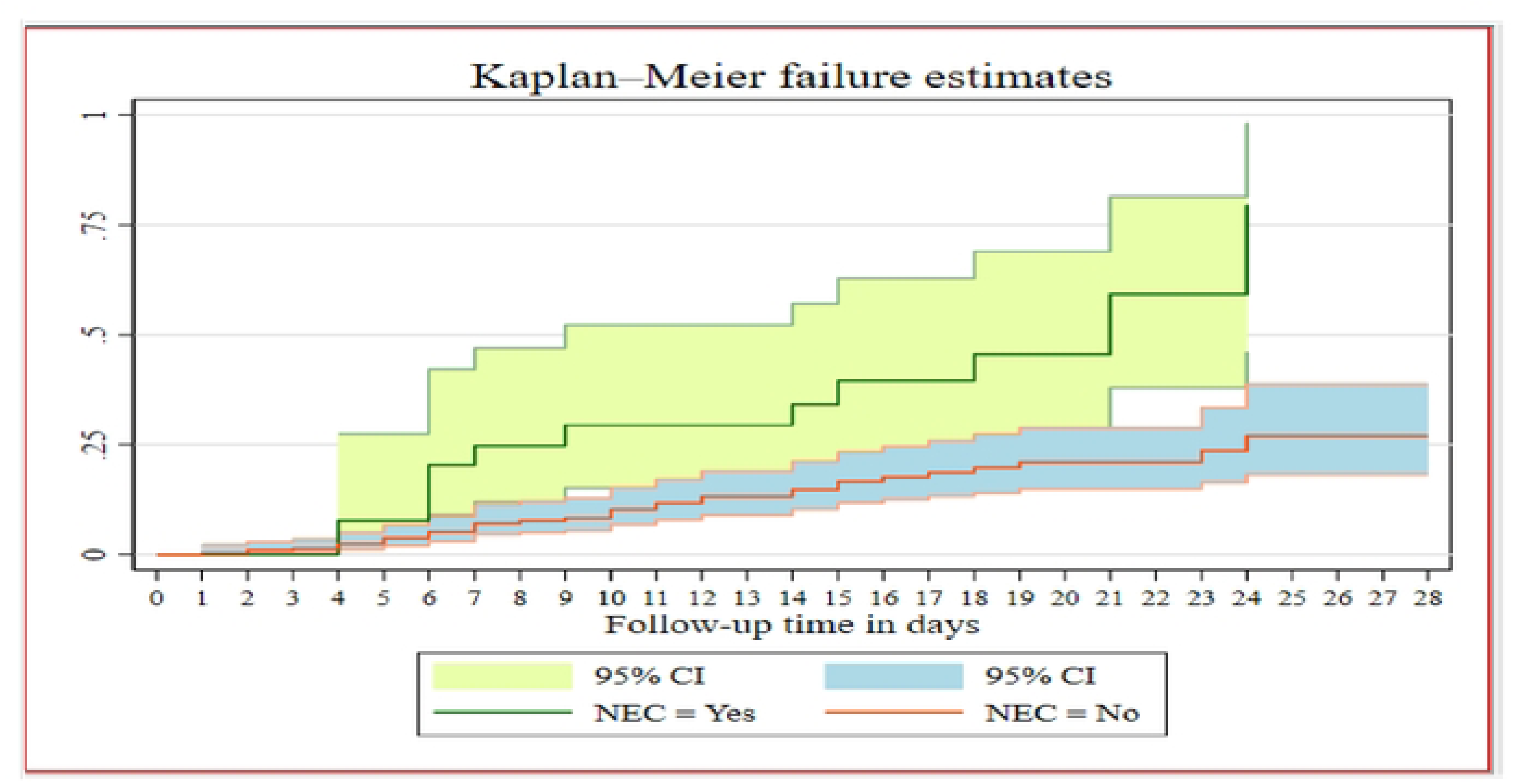
KM failure difference among NEC versus non-NEC thrombocytopenic neonatal death in public hospitals Addis Ababa, Ethiopia, 2025.

**Figure 6:**
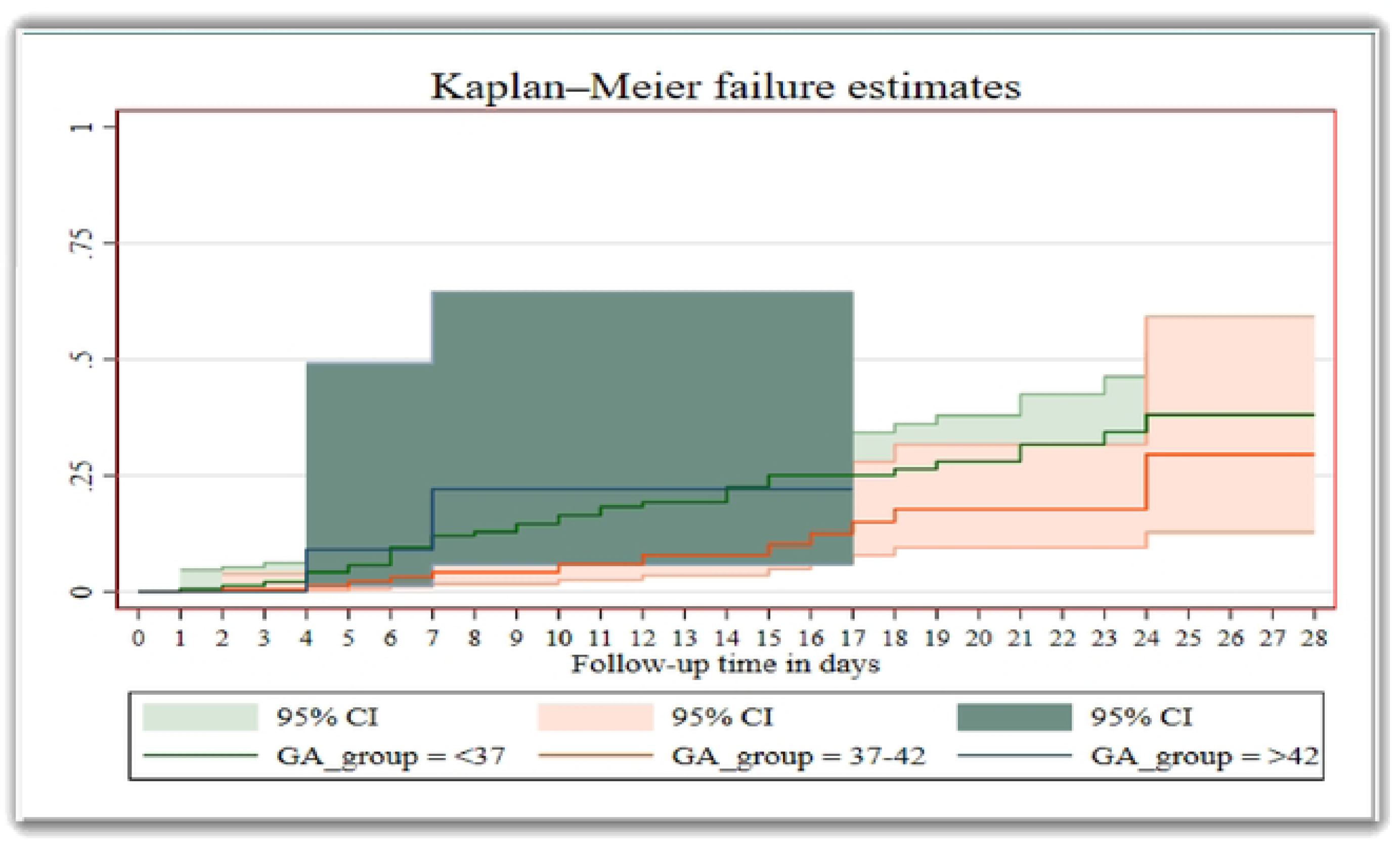
KM failure difference based on categories of gestational age NEC thro1nbocytopenic neonatal death in public hospitals Addis Ababa, Ethiopia, 2025.

### Predictors of time to death for different categorical variables

Several factors were found to be statistically significant predictors of mortality among thrombocytopenic neonates admitted to neonatal intensive care units in this study using bi-variable Cox regression analysis. Severe preeclampsia, prolonged rupture of membranes (PROM), admission age <1 hour, birth weight, perinatal asphyxia (PNA), early-onset neonatal sepsis (EONS), respiratory distress (RD), hospital-acquired infections (HAI), necrotizing enterocolitis (NEC), neonatal hyperbilirubinemia (NHB), delayed feeding initiation, apnea, absence of neonatal nursing round practice, average number of daily visits by pediatricians and neonatologists, and interruptions in CBC machine functionality were all significantly associated to higher mortality.

After bi-variable cox regression, variables having p-value less than 0.25 were transported to multi-variable cox regression. Neonates born to mothers with severe preeclampsia had a significantly higher risk of death. Specifically, thrombocytopenic neonates born from mother having sever eclampsia have 3.8 times higher (AHR = 3.84; 95% CI: 1.78–8.26; p = 0.001) compared with their counter groups.

Similarly, very low birth weight (<1499 grams) was an identified predictor of mortality which increases the hazard of mortality by 3.7 times compared with neonate’s normal birth weight (AHR = 3.67; 95% CI: 1.14–11.80; p = 0.029).

The presence of perinatal asphyxia (PNA) also significantly increased the hazard of death. The hazard of deaths among thrombocytopenic neonates with PNA were 2.8 higher compared to those without PNA (AHR = 2.76; 95% CI: 1.32–5.79; p = 0.004).

Necrotizing enterocolitis (NEC) was another predictor that increase the hazards of death by more than 2.5 times affected neonates had over twice the hazard of death compared to those without NEC (AHR = 2.45; 95% CI: 1.14–5.31; p = 0.022).

Delayed initiation of feeding, defined as starting after 24 hours of birth, was significantly associated with increased hazards of thrombocytopenic mortality. Reveling that thrombocytopenic neonates who didn’t start trophic feeding within the first 24 hours have 3.4 more hazards of death than those who were fed within the first 24 hours (AHR = 3.37; 95% CI: 1.10–10.29; p = 0.033). (Table 7)

**Table 7:**
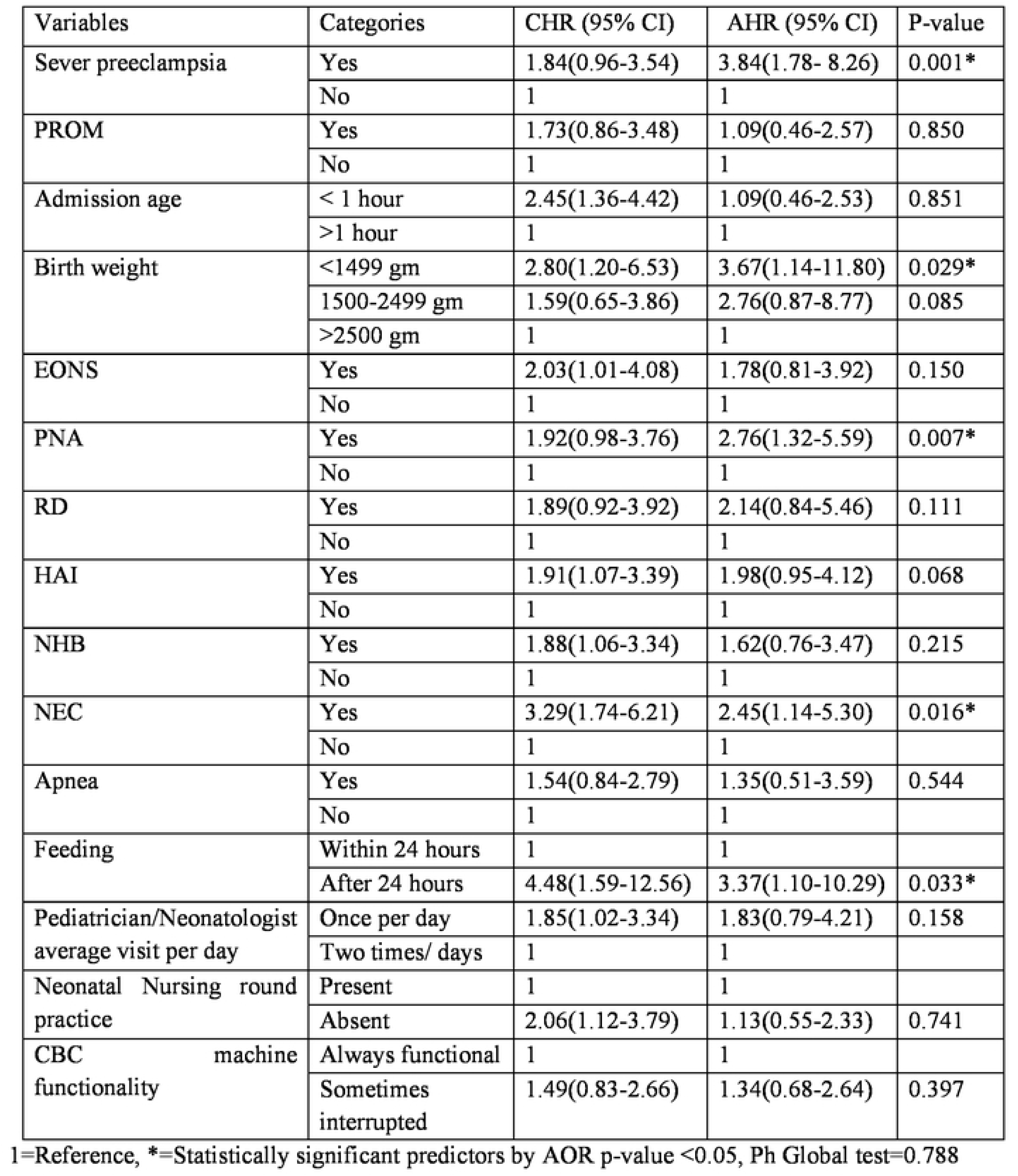
Multi-variable cox proportional hazard regression results for thrombocytopenic neonates admitted to NICU of Addis Ababa public hospitals, Ethiopia, 2025.

For survival analysis, the Cox regression model was first tested. The Cox proportional hazards model was found to be suitable for the dataset based on the model fitness test. The model’s suitability was confirmed by the global test for proportional hazards assumption, which is a p-value of 0.7882, greater than 0.05, indicating no destruction of the assumption.

## DISCUSSION

According to this study the overall mortality of thrombocytopenic neonates admitted to public hospitals of Addis Ababa during the study period was 49(14.1%) (CI:10.4-18.1). This result was consistent to the findings of studies conducted in Egypt 17.24%(36), and India 16.6%(37). The possible explanation for this similarity may be attributed to comparable clinical profiles, including the severity of illness and common causes such as neonatal sepsis. Additionally, similarities in study design, study populations, and operational definitions may have contributed to these consistent findings.

However, the finding of this study was lower than the study conducted Ireland 22.4%(38), and China (26.8%)(39). This discrepancy might be attributed to methodological differences, including variations in study population, study design, sample size, duration of follow-up, and the inclusion and exclusion criteria of neonates. Additionally, discrepancies could be explained by variations in the baseline characteristics of the neonates, such as gestational age, birth weight, and the severity of thrombocytopenia.

This finding was higher than the result found in a studies conducted Nepal (5.40%) (40), and Austria (10.8%)(8). This discrepancy may be due to difference in health care setting may also have an effect where this study was conducted in resource limited settings compared to others studies where thrombocytopenia neonatal survival may be enhanced by countries having more advanced health care settings. Another explanation variations in healthcare infrastructure, availability of timely interventions, and differences in study design or follow-up duration may explain the observed discrepancy.

The incidence density rate of thrombocytopenic neonatal death in this study was 13.04/1000 neonate-day observation. This finding is lower than the neonatal mortality incidence rate reported in a multicenter study conducted in Sub-Saharan Africa, where an overall neonatal mortality IDR of 24.5 per 1000 neonate-days was documented. This may be due to the fact that the current study specifically targeted thrombocytopenic neonates while excluding congenital neonatal conditions, which are often associated with higher mortality. Additionally, the study setting in Addis Ababa may benefit from better healthcare infrastructure, more experienced neonatal staff, and earlier diagnosis and management of thrombocytopenia. Differences in study design, inclusion criteria, and the severity of cases could also contribute to the lower incidence density rate observed in this study. This finding is higher than the neonatal mortality incidence rate reported in Australia, which was 1.8 per 1,000 live births (16). This difference may be explained by the higher burden of comorbidities like sepsis, NEC, and IVH in low-resource settings, where prompt diagnosis and advanced neonatal care are frequently limited, may account for this discrepancy. High-income nations like Australia, on the other hand, have NICUs that are well-equipped, trained staff, and standardized management procedures, all of which help to reduce the mortality rates of high-risk neonates.

The restricted mean time to death in this study was 23.36 days (95% CI: 22.23–24.50), with a standard error of 0.58019. This finding is supported with previous studies conducted in Australia (10.2 days)(16), and Saudi Arbia (18.5 days)(35). This discrepancy might result from delayed hospitalization, extended supportive care initiatives, disparities in case severity or inclusion criteria, or restricted access to advanced neonatal care in Ethiopia. On the other hand, high-income nations frequently have instant access to specialized care, which allows for quicker stabilization or, in certain situations, earlier identification of medical futility and the start of palliative care, which shortens the time to death for critically ill neonates.

In this study, being born from mother having sever preeclampsia was predictors of time to death of thrombocytopenic neonatal mortality which increases the hazard by 3.8 times (AHR: 3.84) compared to their counter parts. This finding is consistent with studies conducted in Egypt(41) India(42), and China (43). This is supported by the fact that severe preeclampsia can make the placenta less effective at getting blood to the baby, which can cause fetal hypoxia, intrauterine growth restriction (IUGR), and preterm birth. All of these things raise the risk of thrombocytopenia and bad outcomes for the baby. Also, preeclampsia can cause problems with the mother’s endothelial cells and inflammation, which can make it harder for the fetus to make platelets and stay alive, which raises the risk of early neonatal death(44).

In this study the neonates born at very low birth weight (<1499 grams) was predictors of time to death of thrombocytopenic neonatal mortality which increases the hazard by 3.7 times (AHR: 3.67) compared to birth weight between 1500–2499 grams. This finding is supported by previous studies conducted in Egypt(45), India(12), and China(39). This could be that neonates born with very low birth weight (VLBW) are highly vulnerable to adverse outcomes due to the immaturity of multiple organ systems. When this physiological immaturity is compounded by thrombocytopenia, the risk of early neonatal death significantly increases(46).

Thrombocytopenic neonates developing perinatal asphyxia (PNA) during the follow up period were 2.8 times (AHR: 2.76) fasten the time to death compared to neonate who didn’t develop perinatal asphyxia (PNA). This was similar with previous studies conducted in Nigeria(47), Netherlands(48), Pakistan(9), and India(49). This is supported by the scientific evidence that perinatal asphyxia makes it hard for vital organs to get enough oxygen, which causes multiple organ dysfunction and a lot of stress on the baby’s body. Thrombocytopenic neonates have a lower platelet count, which makes it harder for their blood to form clots. This raises the risk of bleeding problems like intracranial hemorrhage. PNA-induced inflammation and oxidative stress can also make platelets break down and stop working, which makes thrombocytopenia even worse. This chain of events greatly increases the risk of death in these weak newborns (50).

Necrotizing enterocolitis (NEC) was one of the significant predictors identified in this study which shorten the hazard of time to death by 2.5 times (AHR: 2.45). This finding was almost similar with study done in Austria(8), Iran(51), and Indonesia (52).This is supported by the scientific evidence that necrotizing enterocolitis (NEC) is a serious inflammatory disease of the newborn intestine that kills intestinal tissue and can lead to sepsis, or a systemic infection. When a newborn has thrombocytopenia, their low platelet count makes it harder for clots to form, which raises the risk of bleeding problems in the damaged intestinal tissue. Severe damage to the intestines, inflammation, and bleeding can quickly lead to multi-organ failure and septic shock, which greatly raises the risk of early neonatal death. Systemic inflammation caused by NEC can also make thrombocytopenia worse by destroying and using up more platelets, which speeds up death in a vicious cycle (53).

According to the current study, thrombocytopenic neonates with delayed initiation of feeding, were 3.4 times (AHR = 3.37) more likely to die than those who were fed within the first 24 hours. This is supported by the scientific evidence that neonates who are thrombocytopenic, delayed feeding initiation can hinder gut maturation and jeopardize the intestinal mucosal barrier’s integrity. Early enteral feeding boosts immunological response, encourages the growth of beneficial gut flora, and increases gastrointestinal motility. Neonates who experience delayed feeding are more susceptible to infections like necrotizing enterocolitis (NEC), intestinal atrophy, and bacterial translocation. These risks are increased in neonates with thrombocytopenic disorder, who already have compromised immune systems and hemostasis, making them more vulnerable to systemic infections and complications that raise the risk of death (54).

### Limitation of the study

The study was conducted over a three-month period, which may not capture seasonal variations in neonatal illness patterns and outcomes. Variables such as maternal infections, and socio-economic factors were not included, which may have influenced neonatal outcomes. Additionally, institutional variability was not considered, which may have influenced differences in clinical practices, resource availability, or patient outcomes across study sites.

## Conclusions

Several significant predictors of mortality were identified, including severe preeclampsia, very low birth weight, perinatal asphyxia, necrotizing enterocolitis, and delayed initiation of feeding. Develop standardized protocols for early diagnosis, monitoring, and treatment of thrombocytopenia and its complications such as NEC and PNA. Conduct longitudinal studies to assess long-term outcomes of neonates with thrombocytopenia, including neurodevelopmental and hematological consequences.

## ABBREVIATIONS AND ACRONYMS

AGA: Appropriate For Gestational Age
APGAR: Appearance, Pulse, Grimace, Activity, Respiration
C/S: Cesarean Section
CPAP: Continuous Positive Airway Pressure
DM: Diabetes Mellitus
EONS: Early onset Neonatal Sepsis
HAI: Hospital Acquired Infection
HIV/AIDS: Human Immune Virus /Acquired Immuno Deficiency Syndrome
KMC: Kangaroo Mother Care
LGA: Large For Gestational Age
NEC: Necrotizing enterocolitis
NHB: Neonatal Hyper Bilirubinemia
NICU: Neonatal Intensive Care Unit
PNA: Perinatal Asphyxia
RD: Respiratory Distress
SGA: Small For Gestational Age
SVD: Spontaneous Vaginal Delivery
WHO: World Health Organization

## DECLARATIONS

### Ethics approval and consent to participate

A letter of ethical clearance was first obtained from Menelik II Medical and Health Science College and Research and Community Service Directorate and from Addis Ababa Public Health and Emergency Management Directorate with ethical approval letter reference number of አጠ5/38/248 and አ/አ/ጤ/2/245/17 respectively. Then, an official letter was submitted to the selected study area. Informed consent was taken from the respective index mother or father or other relatives found nearby to the neonate. While the informed consent was taken, the indexed mother and father or relatives were informed as the papers were published in international journals. The neonate’s name and medical record identification information wouldn’t be collected, and confidentiality would be maintained. All data collected from the chart were kept strictly confidential and used only for the study purpose.

### Consent for publication

Not applicable

### Data Availability

All data can be accessed upon request from first author.

### Author Contributions

**Conceptualization:** Yohannes Godie Ashebir, Fikirte Kassaye, Teklu Assefa, Taddle Abate, Tiruye Menshaw, Misrak Tafese, Misgana Hirpha, Asalef Endazanaw

**Data collection:** Asalef Endazanaw, Misgana Hirpha, **Data export and cleaning:** Yohannes Godie Ashebir **Formal analysis:** Yohannes Godie Ashebir

**Investigation:** Yohannes Godie Ashebir, Fikirte Kassaye, Teklu Assefa, Taddle Abate **Methodology:** Yohannes Godie Ashebir, Fikirte Kassaye, Teklu Assefa, Taddle Abate, Tiruye Menshaw, Misrak Tafese, Misgana Hirpha, Asalef Endazanaw

**Software:** Yohannes Godie Ashebir, Fikirte Kassaye, Teklu Assefa, Taddle Abate, Tiruye Menshaw, Misrak Tafese, Misgana Hirpha, Asalef Endazanaw,

**Supervision:** Yohannes Godie Ashebir, Taddle Abate

**Validation:** Yohannes Godie Ashebir,

**Visualization:** Yohannes Godie Ashebir,

**Writing up:** Yohannes Godie Ashebir, Fikerte Kassaye, Teklu Assefa, Taddle Abate, Tiruye Menshaw, Misrak Tafese, Misgana Hirpha, Asalef Endazanaw

### Data Availability

All data can be accessed upon request from first author.

## Acknowledgment

We would like to prompt our genuine gratitude to Menelik II medical and health sciences college. Our sincere appreciation goes to all of the study participants with their indexed mothers who voluntarily consented to agree take part in this study. We also want to express our gratitude to the supervisors and data collectors at Addis Ababa public hospitals that took part for their invaluable assistance and dedication during data collection period.

